# The Effect of a Simple Phone Call Intervention on FIT-Positive Individuals: An Exploratory Study

**DOI:** 10.1101/2020.05.11.20097899

**Authors:** Gretel Jianlin Wong, Jerrald Lau, Ker-Kan Tan

## Abstract

**Purpose:** Colorectal cancer (CRC) screening has been shown to improve patient outcomes. A widely utilised preliminary screening tool is the Faecal Immunochemical Test (FIT). However, follow-up rates after a positive FIT result remain suboptimal.

**Methods:** In order to improve FIT-positive individuals’ compliance to follow-up consultation and to elucidate barriers to action, a simple 5- to 10-minute telephone intervention with a structured script based on the Protection Motivation Theory (PMT) was conducted.

**Results:** Ninety-two FIT-positive individuals who had previously rejected follow-up consultation in the National University Hospital, Singapore were interviewed by the study team. Individuals reported barriers to action such as the denial of a positive FIT result (41.8%) and a lack of knowledge (34.5%). More than 20% of the participants who had yet to schedule follow-up consultation changed their minds after the intervention.

**Conclusion:** The results suggest that a simple, structured telephone call can potentially encourage more FIT-positive individuals to pursue follow-up investigation.

## Introduction

Colorectal cancer (CRC) is the third most common cancer worldwide [1]. CRC screening has been shown to improve patients’ outcomes through earlier detection and treatment of adenomatous polyps and CRC [2].

Singapore has instituted a national CRC screening program using the faecal immunochemical test (FIT) kit, which is free-of-charge to all residents aged 50 years and above [3]. FIT uptake remains suboptimal [4] and most efforts have been focused on increasing uptake rates. However, what is not often highlighted is that there is a significant proportion of individuals who were tested positive for FIT but chose not to undergo subsequent consultation and colonoscopy [5,6].

Literature has suggested that interventions are more often successful when underpinned by a health behaviour theory or framework [7]. The Protection Motivation Theory (PMT) states that individuals make decisions to protect themselves (e.g. perform a health behaviour) through an appraisal of threat (e.g. severity of consequences from not performing the behaviour) and an appraisal of coping (e.g. perceived self-efficacy in ability to perform the recommended behaviour) [8]. PMT has been used across various health related fields to study motivations and behaviours of individuals [9] and has been shown to predict screening outcomes in different cancers [10].

While recent studies have attempted to examine the behavioural factors associated with post-FIT colonoscopy, most interventions to improve compliance have so far been atheoretical [11]. This pilot study was therefore conducted to determine whether a simple intervention based on PMT may be useful in improving the compliance of individuals who initially rejected a follow-up consultation. In addition, common barriers preventing these individuals from seeking follow-up consultation were also collected and elucidated for further understanding.

## Methods

Currently in Singapore, individuals who test positive for FIT via the national screening programme will be contacted by a dedicated coordinator in the restructured hospital nearest to their residential address. The coordinator will arrange for a follow-up appointment with a gastrointestinal (GI) specialist who will then recommend a diagnostic colonoscopy if indicated.

### Ethical Approval

This study was ethically approved by the National Healthcare Group Domain Specific Review Board (NHG DSRB Ref.: 2017/00409). Verbal consent was obtained from all participants.

### Participants

In 2016, the coordinator at the National University Hospital (NUH) Singapore called a total of 1152 individuals who tested FIT-positive. Of those, only 649 (56.3%) agreed to a follow-up consultation with the GI specialist. To test out our hypothesis, we randomly selected 100 FIT-positive individuals who had rejected the coordinator’s arrangement for an appointment from the cohort of patients that were tested positive from 2016 to 2018. These individuals were contacted between February 2018 and January 2019. Of these 100 individuals, 92 (92.0%) agreed to participate in our study.

### Measures and Procedure

Participants were contacted via telephone and a 5- to 10-minute informal interview was conducted by the study team using a standardised telephone script. The script was designed to explore the reasons of non-compliance amongst these individuals, and to then encourage them to schedule and attend a follow-up consultation with a GI specialist. The script utilised facets of PMT by emphasising that further investigations such as colonoscopy would be diagnostic and present a definitive result (coping appraisal), and that these individuals were at an increased risk of CRC due to their FIT-positive status (threat appraisal). The study team addressed participant queries and reiterated the importance of compliance to a follow-up consultation and test. The study team also recorded barriers reported by participants during the interview, and challenged misconceptions presented by the participant, in order to increase the participants’ coping and threat appraisal to encourage attendance of a follow-up consultation. If the participant was willing to go for further investigations, an appointment with a GI specialist at a public institution of their choosing was arranged.

## Results

Majority of the participants were male (59.8%), with a median age of 65 years (range = 60 – 88 years). Forty one of the 92 participants (44.6%) were contacted within six months of the initial notification of FIT-positive status by the coordinator, with the rest being contacted more than six months after initial notification. The median time from the participant being informed by the coordinator of a positive FIT result to the participant receiving a call from the study team was 48.3 weeks (range = 15.0 – 123.0 weeks).

Of the 92 participants, 37 (40.2%) reported that they had already received medical consultation in either a different restructured hospital or a private hospital, and were termed as “doers”. There were two recurring reasons given by doers for seeking alternative arrangements. The first was a desire for an earlier appointment date than the one given by the coordinator. The second was the preference of another institution over the one allocated based on residential proximity.

Several barriers precluded consultation for the remaining 55 participants (59.8%) termed as the “non-doers”. This included 1) denial of the positive FIT result, 2) a lack of knowledge over what a positive FIT result meant, 3) unwillingness to follow-up with an unfamiliar doctor, 4) failure to receive any additional follow-up reminders, 5) perceived inconvenience of an additional medical appointment, 6) fear of procedure and results, 7) high cost for follow-up consultations and procedures, and lastly 8) deeming a follow-up unnecessary due to old age. A descriptive list of barriers can be found in Table 1.

**Table 1:**
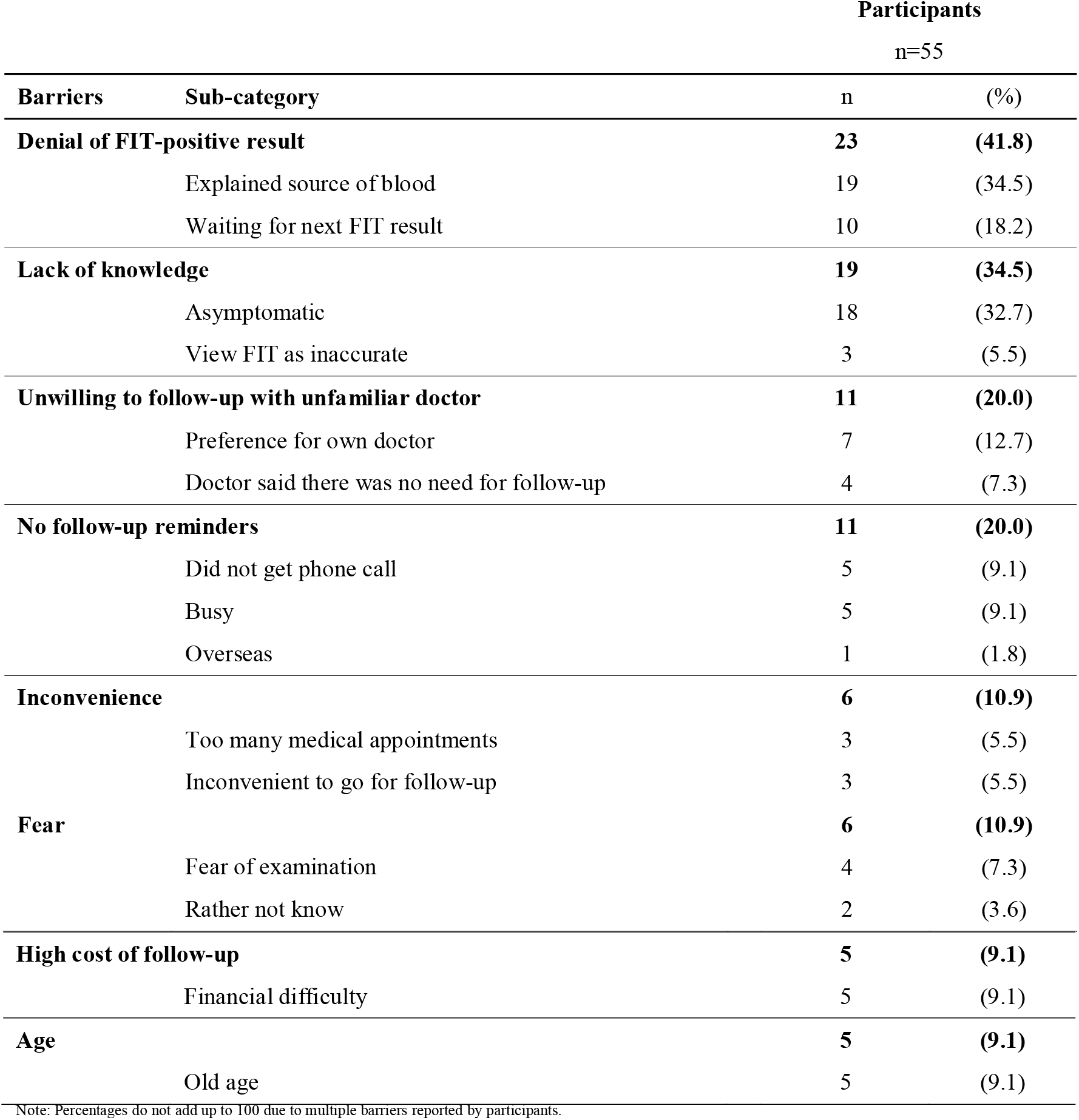
Frequency of barriers to follow-up consultation as reported by non-doers.

Twelve out of 55 non-doers (21.8%) agreed to see a GI specialist after the study team called them. Six non-doers (10.9%) had previously gone through colonoscopy five to 20 years prior, but only one out of the six of them was willing to go for further consultation. The rest rejected consultation, wanting only to do further investigations if their own doctors recommended it. All five non-doers (9%) who reported not receiving follow-up calls were agreeable to further investigations.

## Discussion

Our study showed that a simple phone call could convert over 20% of individuals who initially rejected a follow-up consultation to change their mind. This has significant ramifications for all coordinator-led screening programmes. Individuals with a lack of knowledge may hold the mistaken assumption that a positive FIT result is synonymous with having CRC, and thus, receiving the call relaying the news of a positive FIT result will result in the initial response of shock and denial and hence refusal to seek further consultation. This is demonstrated clearly from our findings as the most frequent barrier reported by non-doers was the denial of a FIT positive result. Many gave reasons for the presence of blood in their stools in order to justify their decision to not go through with a follow-up consultation, wanting to repeat FIT to seek confirmation of a negative result.

Given the sudden nature of the phone call by the coordinator, individuals may need time to accept the implications of a positive test, discuss their results with their loved ones or their general practitioners and explore their options before being open to changing their minds. Thus, what is implicit is that an initial rejection to a referral to see a specialist does not necessarily imply that the patients will not change their minds in future.

Many individuals also appeared to have used the FIT kits without being fully aware of its purpose and implications. Thus, upon receiving the positive test results, some sought to negate its significance or hung on to hope for a negative test as they were not keen to seek further investigations regardless of the FIT results. In fact, it would not be surprising if some patients were in fact symptomatic and did the FIT kits only to allay their fears of harbouring CRC, instead of seeking the definitive opinion of a GI specialist. Thankfully, these reported barriers can be overcome with targeted intervention such as public education, better access, and further subsidy of subsequent investigations and treatment [12].

This study also highlighted the limitation of the current system in reaching out to individuals, as observed by the 9.1% of participants who reported no contact from the coordinator due to their busy schedules or being overseas. A simple follow-up phone call by the study team was sufficient in convincing these participants to schedule a follow-up consultation.

Nonetheless, the small sample size caveats the generalisability of these findings, and participants who were agreeable to participate in this study may not have necessarily been entirely representative of the FIT-positive population at large. However, this study aims to serve as a snapshot of using a simple and quick intervention with a PMT based script to increase participation in further investigation for CRC, as well as to present barriers precluding the individuals from pursuing further investigations.

## Conclusion

To conclude, this exploratory study suggests that a simple telephone intervention can potentially change the minds of FIT-positive individuals who initially rejected a follow-up consultation in the first instance, improving their compliance rates to completion of the CRC screening process. Future studies may wish to examine the effectiveness of such telephone interventions more rigorously through larger cross-sectional studies or community trials, first to investigate the generalisability of the barriers reported, and then to design interventions that target these relevant barriers preventing non-doers from follow-up investigations.

## Data Availability

The data is a result of the MOU between SCS and the PI

## Acknowledgements

We acknowledge Ms Lim Yi Xuan for her contributions to participant recruitment and data collection.

## References

1. Bray F, Ferlay J, Soerjomataram I, Siegel RL, Torre LA, Jemal A (2018) Global cancer statistics 2018: GLOBOCAN estimates of incidence and mortality worldwide for 36 cancers in 185 countries. CA Cancer J Clin 68 (6):394–424. doi:10.3322/caac.21492

2. Citarda F, Tomaselli G, Capocaccia R, Barcherini S, Crespi M (2001) Efficacy in standard clinical practice of colonoscopic polypectomy in reducing colorectal cancer incidence. Gut 48 (6):812–815. doi:10.1136/gut.48.6.812

3. Singapore Cancer Society (n.d.) FIT Kit. https://www.singaporecancersociety.org.sg/get-screened/colorectal-cancer/fit-kit.html.

4. Ministry of Health Singapore (2010) National Health Survey 2010.

5. Tan WS, Tang CL, Koo WH (2013) Opportunistic screening for colorectal neoplasia in Singapore using faecal immunochemical occult blood test. Singapore Med J 54 (4):220–223. doi:10.11622/smedj.2013077

6. Gingold-Belfer R, Leibovitzh H, Boltin D, Issa N, Tsadok Perets T, Dickman R, Niv Y (2019) The compliance rate for the second diagnostic evaluation after a positive fecal occult blood test: A systematic review and meta-analysis. United European Gastroenterol J 7 (3):424–448. doi: 10.1177/2050640619828185

7. Viswanath K RB, Glanz K, (2015) Health Behavior: Theory, Research, and Practice. Fifth edn. Jossey-Bass, San Francisco, CA

8. Rogers RW (1975) A Protection Motivation Theory of Fear Appeals and Attitude Change1. The Journal of Psychology 91 (1):93–114. doi:10.1080/00223980.1975.9915803

9. Floyd DL, Prentice-Dunn S, Rogers RW (2000) A meta-analysis of research on protection motivation theory. Journal of Applied Social Psychology 30 (2):407–429. doi:10.1111/j.1559-1816.2000.tb02323.x

10. Malmir S, Barati M, Khani Jeihooni A, Bashirian S, Hazavehei SMM (2018) Effect of an Educational Intervention Based on Protection Motivation Theory on Preventing Cervical Cancer among Marginalized Women in West Iran. Asian Pac J Cancer Prev 19 (3):755–761. doi:10.22034/apjcp.2018.19.3.755

11. Selby K, Baumgartner C, Levin TR, Doubeni CA, Zauber AG, Schottinger J, Jensen CD, Lee JK, Corley DA (2017) Interventions to Improve Follow-up of Positive Results on Fecal Blood Tests: A Systematic Review. Ann Intern Med 167 (8):565–575. doi:10.7326/m17-1361

12. Community Preventive Services Task Force (2016) Cancer Screening: Multicomponent Interventions - Colorectal Cancer, https://www.thecommunityguide.org/findings/cancer-screening-multicomponent-interventions-colorectal-cancer.

